# The impact of the Covid-19 pandemic on emergency and elective hip surgeries in Norway

**DOI:** 10.1101/2020.09.11.20191734

**Authors:** Karin Magnusson, Jon Helgeland, Mari Grøsland, Kjetil Telle

**Affiliations:** Norwegian Institute of Public Health, Cluster for Health Services Research; Lund University, Faculty of Medicine, Department of Clinical Sciences Lund, Orthopaedics, Clinical Epidemiology Unit, Lund, Sweden

## Abstract

**Objective:** To assess the effects of Covid-19 pandemic lockdown restrictions on the number of emergency and elective hip joint surgeries, and explore whether these procedures are more/less affected by lockdown restrictions than other hospital care.

**Methods:** In 1.344.355 persons aged ≥35 years in the Norwegian emergency preparedness (BEREDT C19) register, we studied the daily number of persons having 1) emergency surgeries due to hip fractures, and 2) electively planned surgeries due to hip osteoarthritis before and after Covid-19 lockdown restrictions were implemented nationally on March 13^th^ 2020, for different age and sex groups. Incidence Rate Ratios [IRR] reflect the after-lockdown number of surgeries divided by the before-lockdown number of surgeries.

**Results:** After-lockdown elective hip surgeries were one third the number of before-lockdown (IRR ∼0.3), which is a greater drop than the drop seen in all-cause elective hospital care (IRR ∼0.6) (no age/sex differences). Men aged 35-69 had half the number of emergency hip fracture surgeries (IRR ∼0.6), whereas women aged ≥70 had the same number of emergency hip fracture surgeries after lockdown (IRR ∼1). Only women aged 35–69 and men aged ≥70 had emergency hip fracture surgery rates after lockdown comparable to what may be expected based on analyses of all-cause acute care (IRR ∼0.80)

**Conclusion:** Important to note for future pandemics management is that lockdown restrictions may impact more on scheduled joint surgery than other scheduled hospital care. Lockdown may also impact on the number of emergency joint surgeries for men aged ≥35 but not for women aged ≥70.

## Introduction

The coronavirus disease 2019 (Covid-19) emerged in late 2019 in China and rapidly spread in a global pandemic causing more than 300,000 deaths in a few months (1). As a reaction, many countries implemented different “lockdown” policies, in which most societal institutions, including certain departments at hospitals, were closed or operated with reduced capacity. Norway implemented one of the strictest lockdown policies of all countries at an early stage. The lockdown measures are believed to dramatically have limited the spread of the virus in the country, but on the other side, may have had several unknown negative side effects on the planned and acute care for vulnerable groups. As an example, people with osteoporotic fractures and osteoarthritis are often elderly and fragile (2), with a high need for care to prevent long term disability and death. The conditions are often managed by the most commonly performed surgical joint procedure worldwide: total hip joint surgery (3). Hip fractures, for example, require immediate joint surgery and immobilization on the same or next day of the accident and can be fatal if else (4), whereas hip osteoarthritis is a chronic painful joint disorder that can often be treated with physiotherapy and analgesics alone, but requires surgery in severe cases. Surgeries due to hip osteoarthritis are typically planned weeks or months in advance (5).

The impact of the Covid-19 lock down restrictions on such typically acute and elective care for age-related conditions that require hospitalization is currently unknown, but can be hypothesized to be major, at least for elective care. Also, if there is an effect on acute care, knowledge of which population groups to a lesser extent need, or make use of acute care, is important in the future handling of pandemics. Such analyses may also provide a knowledge foundation for future natural experiments evaluating whether any care is in fact unnecessary in a long term perspective (6). Thus, we aimed to assess the effects of Covid-19 pandemic lock down restrictions on acute and elective inpatient care in Norway during spring 2020, using surgeries for hip fractures and surgeries for hip osteoarthritis as examples.

## Methods

We utilized data from the BEREDT C19 register, which is a newly developed emergency preparedness register aiming to provide rapid knowledge of the spread of the Covid-19 virus and how spread as well as measures to limit spread affect the population’s health, use of health care services and health-related behaviors (7). The register currently consists of electronic patient records from all hospitals in Norway (NPR), data from the Norwegian Surveillance System for Communicable Diseases (MSIS) and the Norwegian Intensive Care and Pandemics Register (NIPaR) which are merged on the personal identification number and updated daily, with a range of other registry linkages currently ongoing. The register covers all data from hospitals (inpatient, outpatient and day-care), with complete diagnostic and procedure codes from January 1^st^ 2020 until the pandemic is over and evaluated. In the current study, our population included everyone in Norway as registered with acute or elective inpatient care and we restricted our sample to the age groups to which diagnoses of hip osteoarthritis and fractures apply (age 35 or more). The BEREDT C19 register is authorized by Norwegian law and no external ethical board review was required.

### Outcomes

Besides studying all registered inpatient care coded with emergency grades acute vs. elective (any cause), we studied the number of patients hospitalized with the outcomes 1) emergency surgeries due to hip fractures, and 2) electively planned surgeries due to hip osteoarthritis. Hip fracture surgeries were identified as having ICD-10 codes S72* (main diagnosis or other diagnosis) in combination with NCSP procedure code NFJ* and/or NFB*, and an emergency grade coded as acute. Hip osteoarthritis surgeries were identified as having ICD-10 codes M16* (main diagnosis or other diagnosis) and procedure code NCSP NFB* and an emergency grade coded as elective.

### Statistical analysis

We assessed the described outcomes prior to and after lockdown restrictions were implemented in Norway on March 13^th^ 2020, i.e. in the period of January to May 2020. We first studied all-cause acute and elective inpatient care using a Poisson regression model with the daily number of hospitalizations as outcome (i.e. acute and elective emergency grades in separate analyses) and time as explanatory variables. Thus, we categorized dates in five 2-weeks periods before March 13^th^ 2020, and five 2-weeks periods after March 13^th^ 2020, covering a total time period from January 3^rd^ 2020 to May 21^st^ 2020. To observe trends in number of surgeries over time, we compared the incidence rate ratios (IRR) for all the 2-week periods with the base level, which was defined as the two first weeks of January, starting from January 3^rd^. We also compared IRR in the 10 weeks before and after lockdown restrictions were implemented nationally in Norway on March 13^th^ 2020, using the period of 10 weeks before lockdown as base level (Jan 3^rd^-March 12^th^). The IRR should be interpreted as the observed number of surgeries for the given period divided by the observed number of surgeries in the base level period.

We secondly repeated these analyses for emergency hip fracture surgeries and elective hip osteoarthritis surgeries. Also, to explore whether any hip surgery patient groups may be more affected by lockdown restrictions in terms of their health care use than what could be expected from our analyses of all-cause acute and elective hospitalizations, we stratified the analyses on age (35–69 years vs. 70 and above), for men and women separately. Analyses were adjusted for weekends and holidays. Finally, we predicted number of unperformed surgeries in the period after March 13^th^ 2020 from the same Poisson regression model, conditional on week days. We used Stata version 16.1 (StataCorp) for all analyses.

## Results

BEREDT C19 comprised 1.344.355 persons with at least one contact with specialist care from January 3rd 2020 to May 21^st^ 2020. Of persons aged 35 or more having > 24h hospitalizations (inpatients), we observed 73.091 emergency and 187.714 elective hospitalizations (persons could here be counted in both groups). Figure 1 shows that persons receiving elective inpatient care had half the rate of care after lockdown (n=26.360 hospitalizations) compared to before lockdown (n=46.731 hospitalizations) (IRR=0.56, 95% confidence intervals [CI]=0.56–0.57 as compared to base levels before lockdown, IRR=1). Also, acute care occurred at a 20% lower rate after lockdown (n=83.838 hospitalizations) than before (n=103.875 hospitalizations) (IRR=0.81, 95% CI=0.80–0.81) (Figure 1).

**Figure 1.**
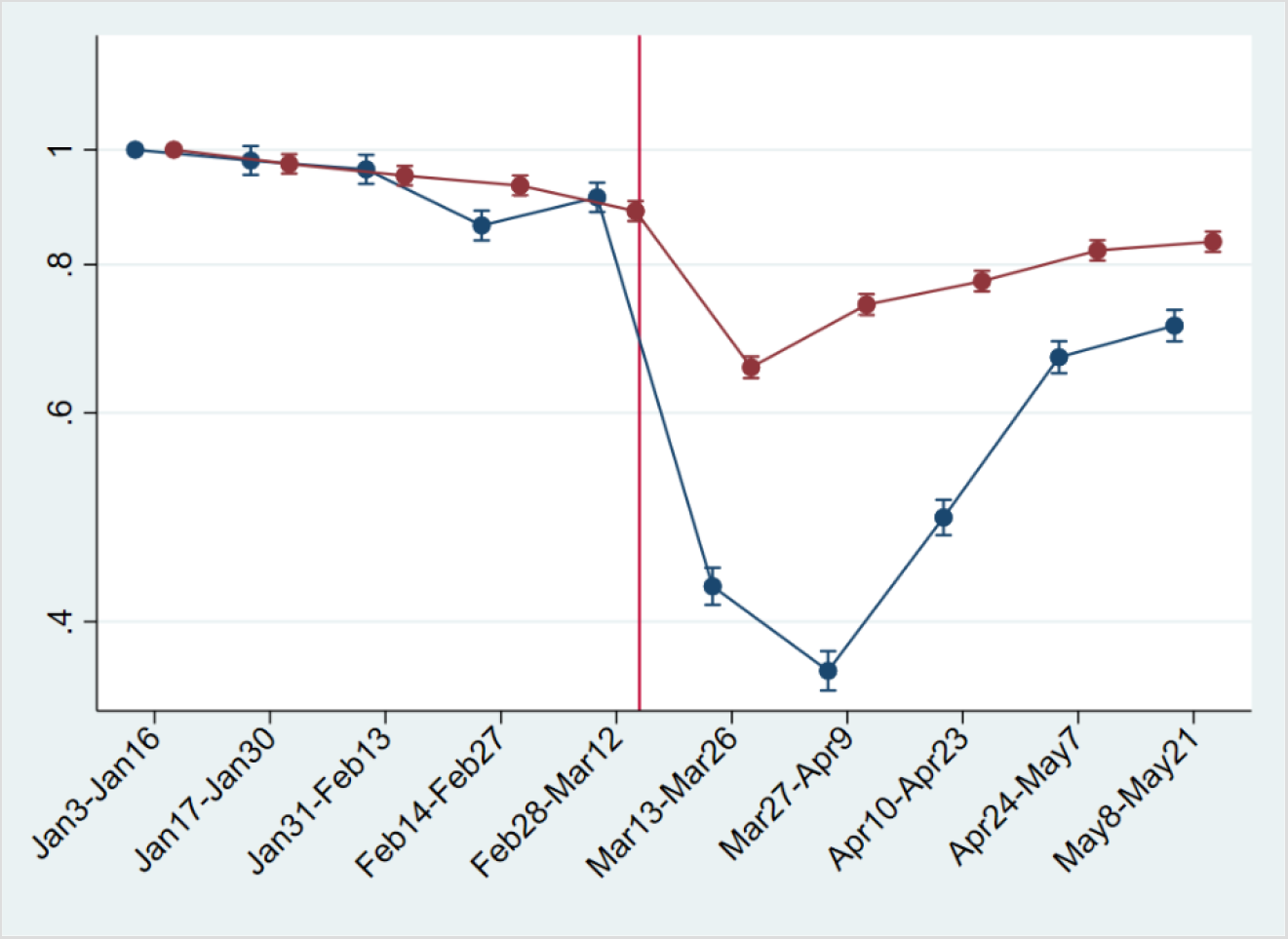
The incidence rate ratios (y-axis, IRR) of daily hospitalizations (any cause) with acute and elective emergency grade in Norway, Jan 3^rd^ 2020-May21st 2020, with Jan 3^rd^ – Jan 16^th^ as base level, with their 95% confidence intervals (CI). Red dots/line = acute care. Blue dots/line = elective care. The red and blue two-weekly dots are graphed next to each other for improved readability. Vertical line represents the national implementation of lockdown strategies on March 13^th^ 2020.

We observed in total 2701 new hip surgeries due to osteoarthritis and 3650 new hip surgeries due to fractures throughout the study period, i.e. in persons without prior prosthesis surgery in any of the hip joints. Before March 13^th^ 2020, the daily number of new hip surgeries corresponded to that reported for the same time period in previous years (8). The rate of elective hip surgeries due to osteoarthritis dropped significantly after lockdown restrictions were implemented on March 13^th^ 2020, with similar observations across age and sex groups (Figure 2). For men aged 35-69, there was also a slight decrease in the rate of emergency hip surgeries due to fractures (Figure 2).

**Figure 2.**
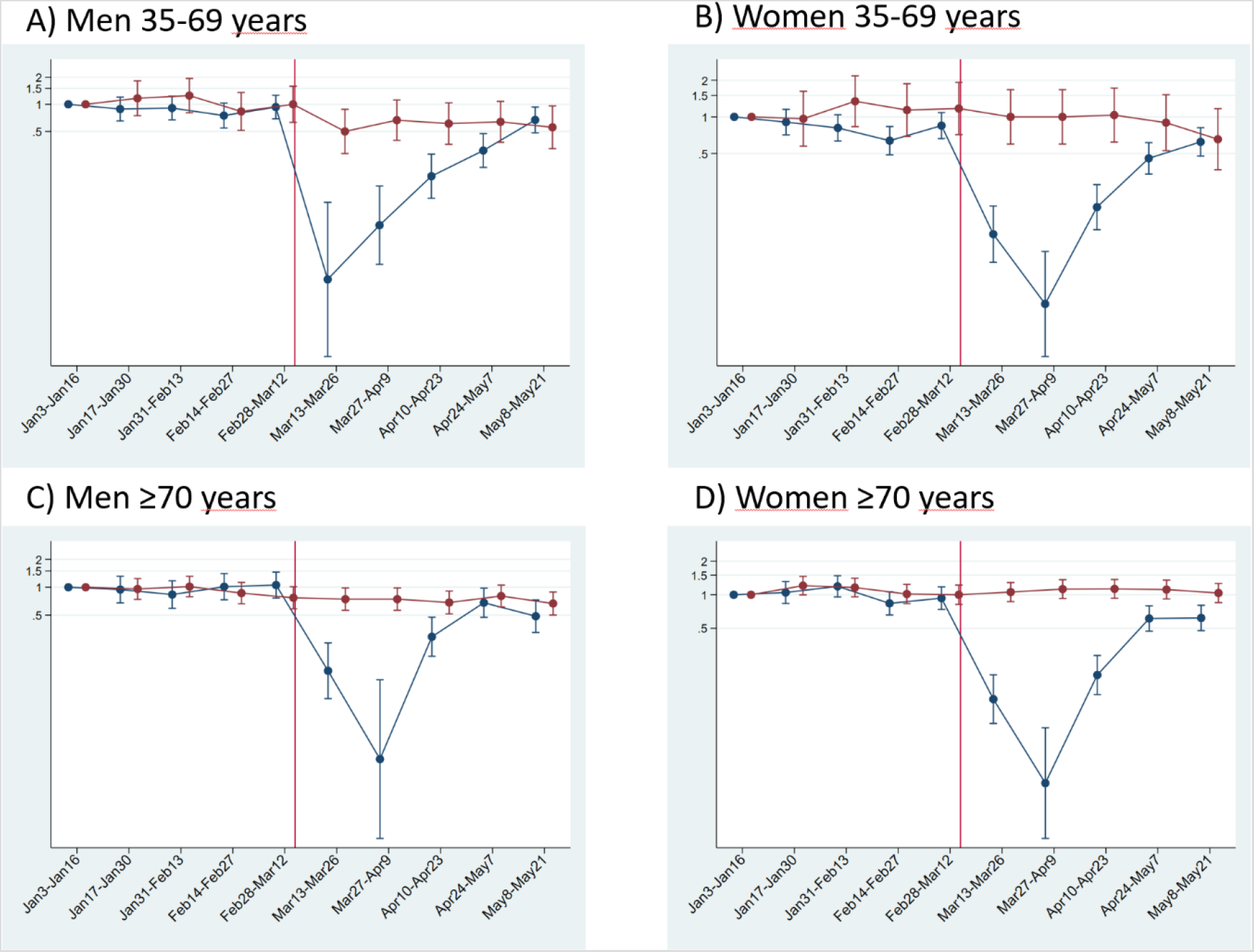
The incidence rate ratios (y-axis, IRR) of emergency hip surgeries due to fracture (red) and the IRR of elective hip surgeries due to osteoarthritis (blue) in Norway, Jan 3^rd^ 2020-May21st 2020 with Jan 3^rd^ – Jan 16 as base level with their 95% confidence intervals (CI). Red dots/line = emergency hip fracture surgeries. Blue dots/line = elective hip osteoarthritis surgeries. The red and blue two-weekly dots are graphed next to each other for improved readability. Vertical line represents the national implementation of lockdown strategies on March 13^th^ 2020.

When compared to what may be expected, based on the average after-lockdown drop in all-cause elective and all-cause acute hospitalizations, we observed large deviations for our musculoskeletal outcomes. For elective hip osteoarthritis surgeries, IRRs were half to that observed for elective all-cause hospitalization, for all age- and sex strata (IRR ∼0.3 for elective hip osteoarthritis surgeries vs. IRR ∼0.6 for all-cause elective care) (Table 1). This would imply that around 1400 planned hip surgeries in Norway have not been performed due to the Covid-10 pandemics and would need to be treated elsewhere or to be scheduled for surgery on another date (estimated no. of unperformed hip osteoarthritis surgeries after March 13^th^ 2020 = 1417 [95% CI]= 1392–1440).

**Table 1:**
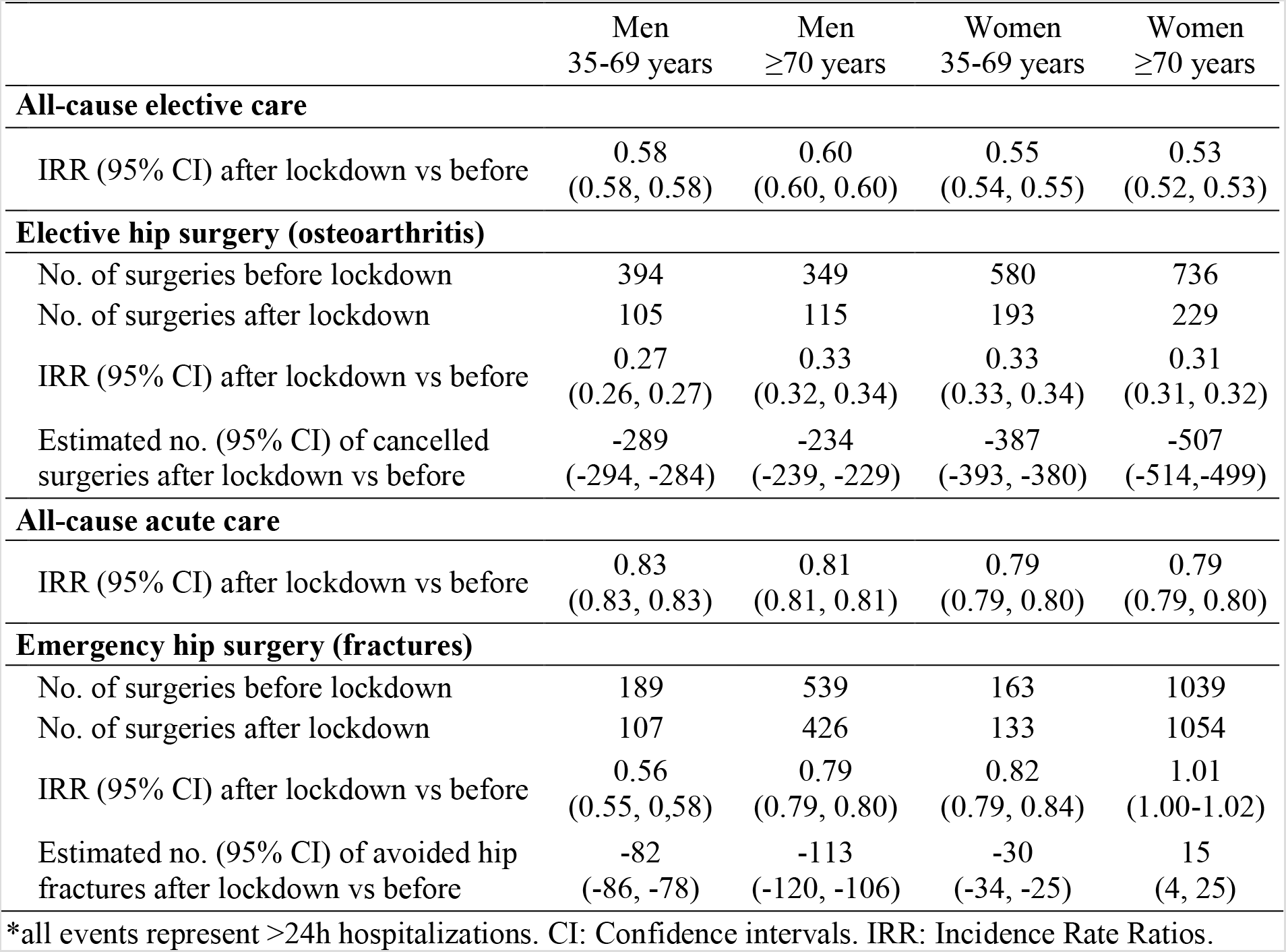
Hip surgeries 10 weeks after compared with 10 weeks before (base level) the implementation of national lockdown on March 13th 2020.

|  | Men<br>35-69 years | Men<br>≥70 years | Women<br>35-69 years | Women<br>≥70 years |
| --- | --- | --- | --- | --- |
| **All-cause elective care** |  |  |  |  |
| IRR (95% CI) after lockdown vs before | 0.58<br>(0.58, 0.58) | 0.60<br>(0.60, 0.60) | 0.55<br>(0.54, 0.55) | 0.53<br>(0.52, 0.53) |
| **Elective hip surgery (osteoarthritis)** |  |  |  |  |
| No. of surgeries before lockdown | 394 | 349 | 580 | 736 |
| No. of surgeries after lockdown | 105 | 115 | 193 | 229 |
| IRR (95% CI) after lockdown vs before | 0.27<br>(0.26, 0.27) | 0.33<br>(0.32, 0.34) | 0.33<br>(0.33, 0.34) | 0.31<br>(0.31, 0.32) |
| Estimated no. (95% CI) of cancelled surgeries after lockdown vs before | -289<br>(-294, -284) | -234<br>(-239, -229) | -387<br>(-393, -380) | -507<br>(-514, -499) |
| **All-cause acute care** |  |  |  |  |
| IRR (95% CI) after lockdown vs before | 0.83<br>(0.83, 0.83) | 0.81<br>(0.81, 0.81) | 0.79<br>(0.79, 0.80) | 0.79<br>(0.79, 0.80) |
| **Emergency hip surgery (fractures)** |  |  |  |  |
| No. of surgeries before lockdown | 189 | 539 | 163 | 1039 |
| No. of surgeries after lockdown | 107 | 426 | 133 | 1054 |
| IRR (95% CI) after lockdown vs before | 0.56<br>(0.55, 0.58) | 0.79<br>(0.79, 0.80) | 0.82<br>(0.79, 0.84) | 1.01<br>(1.00-1.02) |
| Estimated no. (95% CI) of avoided hip fractures after lockdown vs before | -82<br>(-86, -78) | -113<br>(-120, -106) | -30<br>(-34, -25) | 15<br>(4, 25) |

\*all events represent >24h hospitalizations. CI: Confidence intervals. IRR: Incidence Rate Ratios.

For emergency hip fracture surgeries there were large variations by age and sex. Men aged 35-69 had a lower rate of emergency hip fracture surgery than of all-cause acute care (IRR ∼0.6 vs. ∼0.8), whereas women aged 70 or more had a higher such rate (IRR ∼0.8 vs. ∼1.00) (Table 1). In contrast, women aged 35-69 and men aged ≥70 had hip fracture surgery rates comparable to what may be expected based on analyses of all-cause acute care (IRR ∼0.80) (Table 1). Altogether, lockdown restrictions may have given a reduction of ∼200 acute events requiring immediate hip fracture surgery (estimated no. of avoided hip fractures after March 13^th^ 2020 = 210 [95% CI]= 184–236).

## Discussion

In this study of BEREDT C19 - the Norwegian emergency preparedness register - we report a sudden and steep decrease in the daily number of planned hip joint surgeries, beginning on the day after lockdown restrictions were implemented in Norway on March 13^th^ 2020. This decrease was greater than the decrease in other (all-cause) elective inpatient care for all age and sex groups. Interestingly, we also report a consistent decrease of all-cause acute inpatient care that only partly was found in age- and sex-specific analyses of emergency hip joint fracture surgeries: Lockdown restrictions may impact on the number of acute joint surgeries for middle-aged and elderly men (aged ≥35) as well as for middle-aged women (age 35-69), but not for elderly women (aged ≥70).

The observed decrease of elective care including hip surgeries is not surprising, and sheds new light on a recent report on effects of lockdowns on elective care globally (9). Here, we additionally show that the activity of elective surgeries was reduced more than other elective activities in inpatient care, and that it increased rather quickly again, as authorities gained control over the spread of the pandemic during April 2020. However, surgery rates were not back to normal by the end of May 2020 and around 1400 elective hip osteoarthritis surgeries would have to be rescheduled or treated in primary care. Figure 2 shows that the activity recovered approximately equally for the different age and sex groups, although there may be minor variations.

For emergency hip surgeries due to fractures, we observed a somewhat unexpected decrease in incidence that was more evident for men than for women. Hospitals were not instructed to limit access to acute care, so the observed decrease may be explained by the fact that people stayed more at home/inside, which reduced the risk of falls and subsequent hip fractures. If so, the non-decreasing incidence of hip fractures in women may be explained by female hip fractures more frequently being a result of intrinsic causes like bone mineral density (10). Whereas all age and sex groups had fewer hip fracture surgeries after lockdown, women aged ≥70 had a slightly increased hip fracture surgery rate, with an additional 4-25 surgeries occurring in the 10 weeks after lockdown compared to the 10 weeks before lockdown.

Our findings may have important implications for the handling of future new outbreaks of Covid-19. First, our data show that when elective health care and other parts of society are locked down by the authorities, the instructions are followed by the hospitals and the use of elective care decreases to a similar magnitude for men and women, young and elderly. Using hip osteoarthritis surgery as an example, we also show that elective joint surgery rates decrease more after lockdown restrictions are implemented, than other elective inpatient care. Second, and important to note for policy-makers from our study, is that the lockdown restrictions likely also impacted on the need for / use of acute health care, and did so to a different extent for men and women when exemplified using emergency hip fracture surgeries. Here, we cannot distinguish between experiencing an acute event and seeking health care. However, it may be hypothesized that different population groups have different levels of anxiety of seeking health care when a pandemic is present, i.e. people may be afraid of seeking health care because of risk of infection. Because mortality risk is often increased for acute events that are treated, and utterly increased for non-treated acute events, any effort to prevent anxiety of seeking acute care may save lives. In that regard, our findings emphasize the need for correct and targeted information to specific groups of the population, to reduce both pandemic-related anxiety of seeking health care, and to minimize the risk of death along acute events in a period of lockdown.

Some important limitations should be mentioned. First, we could not study whether the effect of lockdown restrictions is causal. For example, it is possible that the decrease in emergency hip fracture surgeries in men aged 35-69 partly are due to seasonal variations, and we suggest this as a topic for future studies. However, we note that the number of surgeries prior to March 13^th^ 2020 was similar to that reported for previous years (8). Also, our findings only apply to Norwegian conditions and countries having similar health care services, health care organization and demography as Norway. Future studies should explore effects of lockdown restrictions on health care use comparing different countries’ lockdown strategies. A second limitation may be that we could not distinguish between experiencing joint pain and/or an acute event and seeking health care. Thus, as described above, there may be age and sex differences in care-seeking behavior that we could not account for here. Finally, there may be several potential competing risks in our sample. For example, persons hospitalized for cancer treatment may be unlikely to experience a hip fracture because they are more indoors. However, our goal was not to study disease etiology, rather, we give an overview of potential impacts persons in need of hip joint surgery may experience as a result of lockdown restrictions.

In conclusion, we show that the lockdown restrictions implemented in Norway due to the spread of Covid-19 pandemics likely reduced the use of elective inpatient care, but also acute inpatient care. Especially, we report that men and midlife age groups had lower rate of emergency hip fracture surgeries after than before lockdown. We believe these findings are important to report for an improved knowledge foundation allowing for an optimal management of future pandemics of a similar or larger scale.

## Data Availability

Data are not available for public use.

## Acknowledgements

We would like to thank the Norwegian Directorate of Health, in particular Director for Health Registries Olav Isak Sjøflot and his department, for excellent cooperation in establishing the emergency preparedness register. We would also like to thank Gutorm Høgåsen and Ragnhild Tønnessen for their invaluable efforts in the work on the register. The interpretation and reporting of the data are the sole responsibility of the authors, and no endorsement by the register is intended or should be inferred. We would also like to thank everyone at the Norwegian Institute of Public Health who has been part of the outbreak investigation and response team.

## References

1. Organization WH. Coronavirus disease (COVID-2019) situation reports.

2. Lombnaes GO, Magnusson K, Osteras N, Nordsletten L, Risberg MA, Hagen KB. Distribution of osteoarthritis in a Norwegian population-based cohort: associations to risk factor profiles and health-related quality of life. Rheumatol Int. 2017;37(9):1541–50.

3. Learmonth ID, Young C, Rorabeck C. The operation of the century: total hip replacement. Lancet. 2007;370(9597):1508–19.

4. Socci AR, Casemyr NE, Leslie MP, Baumgaertner MR. Implant options for the treatment of intertrochanteric fractures of the hip: rationale, evidence, and recommendations. Bone Joint J. 2017;99-b (1):128–33.

5. Zhang W, Moskowitz RW, Nuki G, Abramson S, Altman RD, Arden N, et al. OARSI recommendations for the management of hip and knee osteoarthritis, Part II: OARSI evidence-based, expert consensus guidelines. Osteoarthritis Cartilage. 2008;16(2):137–62.

6. Moynihan R, Johansson M, Maybee A, Lang E, Légaré F. Covid-19: an opportunity to reduce unnecessary healthcare. BMJ. 2020;370:m2752.

7. Norwegian Institute of Public Health. Beredskapsregisteret for covid-19. 2020. https://www.fhi.no/sv/smittsomme-sykdommer/corona/norsk-beredskapsregister-for-covid-19/

8. Helse Bergen HF Ok, Haukeland universitetssykehus. Nasjonalt register for leddproteser. Rapport juni 2014.

9. COVIDSurg ollaborative. Elective surgery cancellations due to the COVID-19 pandemic: global predictive modelling to inform surgical recovery plans. Br J Surg. 2020. https://doi.org/10.1002/bjs.11746

10. Emaus N, Omsland TK, Ahmed LA, Grimnes G, Sneve M, Berntsen GK. Bone mineral density at the hip in Norwegian women and men—prevalence of osteoporosis depends on chosen references: the Tromsø Study. European Journal of Epidemiology. 2009;24(6):321–8.

